# NUCOG10: The development and validation of a short-form of the NUCOG cognitive assessment tool

**DOI:** 10.1101/2025.08.13.25333570

**Authors:** Anna D. Li, Samantha M. Loi, Charles B. Malpas, Justin S.Y. Tu, Dennis Velakoulis, Mark Walterfang

**Affiliations:** Neuropsychiatry Centre, Royal Melbourne Hospital, Melbourne, VIC, Australia; Department of Psychiatry, University of Melbourne, Parkville, VIC, Australia; Melbourne School of Psychological Sciences, Parkville, VIC, Australia; Department of Medicine (Royal Melbourne Hospital), University of Melbourne, Parkville, VIC, Australia; Department of Neurology, Royal Melbourne Hospital, Parkville, VIC, Australia; Florey Institute of Neuroscience and Mental Health, Parkville, VIC, Australia; Department of Health and Medical Sciences, Edith Cowan University, WA, Australia

**Keywords:** Cognitive, dementia, assessment, screening tool

## Abstract

**Objective:** The Neuropsychiatry Unit Cognitive Assessment (NUCOG) is a valid and reliable screening tool used in detecting cognitive deficits in a range of neurological and psychiatric conditions. We aimed to develop abbreviated versions of the NUCOG tool using retrospective data, and to assess their psychometric performance in distinguishing between healthy cognition and dementia.

**Methods:** Healthy controls (*n*=132, 41%) and those with dementia (*n*=191, 59%) were randomised into a ‘training’ cohort (*n*=134, 70%) for the development and a ‘testing’ cohort (*n*=57, 30%) for validation of the short-form versions. Receiver operating characteristic (ROC) curves were first computed for each of the 24 original NUCOG items. Items were ranked according to area under the curve (AUC) values to create 5-item, 10-item and 15-item short-form versions, which were subsequently validated.

**Results:** The psychometric properties of the NUCOG short-form versions were comparable to the original, with all maintaining high convergent validity and reliability. Of the three versions, the 10-item version strikes the ideal balance of breadth and brevity. With a cut-off score of 42/54, the 10-item version generated similar sensitivity, specificity and predictive values for dementia as the original NUCOG, with a sensitivity of 0.98, specificity of 0.95, and positive and negative predictive values of 0.97.

**Conclusions:** The 10-item NUCOG (“NUCOG10”) has strengths in its shorter administration time, of approximately 10 minutes, high reliability and validity, and retention of items from each cognitive domain from the original NUCOG. Future research may involve testing these short-forms in non-tertiary settings, across dementia subtypes and in non-dementia groups.

## Introduction

Cognitive deficits feature in a variety of neuropsychiatric and neurodegenerative disorders, including but not limited to traumatic brain injury, stroke, mild cognitive impairment (MCI), dementia, schizophrenia, bipolar disorder and depression (Cristofori and Levin, 2015; Morozova et al., 2022; Pendlebury and Rothwell, 2009; Rock et al., 2014). Cognitive screening tests are widely used in detecting early cognitive changes and in monitoring for change over time, in a range of settings including medical clinics, on hospital wards and in clinical trials. Each of the widely used screening tools, which include the Mini-Mental State Examination (MMSE) (Folstein et al., 1975), the Montreal Cognitive Assessment (MoCA) (Nasreddine et al., 2005) and the Addenbrooke’s Cognitive Examination (ACE) (Mathuranath et al., 2000), have differing strengths and limitations in certain populations, and vary in breadth and brevity of their cognitive testing, with many short-form versions subsequently produced.

The Neuropsychiatry Unit Cognitive Assessment (NUCOG) tool is a validated and reliable cognitive test that was developed in 2000 within the Neuropsychiatry Unit at the Royal Melbourne Hospital, in the context of assessing complex and comorbid patients who had been referred from tertiary inpatient, outpatient and consultation-liaison service settings (Walterfang et al., 2006). This 24-item tool takes at least 20 minutes to administer, and covers a breadth of cognition, with items organised into five cognitive domains: attention, memory, language, executive and visuo-constructional function. In comparison to the MMSE, it was found to be superior in differentiating dementia patients from controls and in differentiating between dementia subgroups (Walterfang et al., 2006). The NUCOG has been validated in a range of neurological, psychiatric and dementia populations, and translated into different languages including Chinese, Malay and Persian (Barekatain et al., 2010; Chandrasekaran et al., 2010; Gao et al., 2014; Walterfang et al., 2011).

While formal psychometric testing may be required in forming a detailed cognitive profile for patients, it can be time-intensive and not readily available. Short-form cognitive tests offer a solution where rapid and reliable screening at the bedside or in a clinical office setting is possible, especially when time with the patient is limited. In this study, we present the development and validation of three short-form versions of the NUCOG, with varying breadth and time to administer, and assess the reliability, validity and clinical utility of each. The short-form versions were created after ranking each NUCOG item’s ability to discriminate between normal controls and those with dementia. All short-form versions retain the core cognitive domains essential for detecting cognitive deficits in a range of neurological and psychiatric conditions. We aimed to assess the performance of each abbreviated form to determine if they retained the ability to detect dementia in a clinically heterogeneous sample.

## Methods

### Data collection

Data comprised of healthy controls and patients with a principal psychiatric, neurological, dementia or metabolic diagnosis. Control subjects came from the original NUCOG validation study, which included relatives and carers of neurology or neuropsychiatry patients seen in outpatient clinics, university students and staff, and respondents to community advertisements (Walterfang et al., 2006), and from an unpublished control dataset collected in as part of a schizophrenia study conducted at Princess Alexandra Hospital in Brisbane, Australia. For those belonging to the disease group, retrospective clinical data was collected from an online NUCOG database which largely consisted of clinical patients seen at the Neuropsychiatry Unit at the Royal Melbourne Hospital, and from medical records at the same hospital. Data collected included age, gender, rater, years of education, setting, diagnoses, medications, and individual item and global NUCOG scores.

The NUCOG was administered to patients between February 2001 to March 2023 in inpatient and outpatient settings in both public and private facilities in Victoria, Australia. Each NUCOG was undertaken by consultant psychiatrists, registrars or medical officers with affiliations with the Neuropsychiatry Unit at the Royal Melbourne Hospital. Those with missing NUCOG subset scores were removed. Only one NUCOG score was considered per subject; in cases where more than two NUCOGs were available, the one administered closest to the principal diagnosis was retained. Dementia diagnoses were required to have been deemed at least ‘probable’ certainty by a consultant medical practitioner to be included in this study. Diagnostic criteria used was the most contemporaneous, such as the International Working Group 2 (IWG-2) criteria for Alzheimer’s disease (Dubois et al., 2014) and the Rascovsky criteria for behavioural variant frontotemporal dementia (Rascovsky et al., 2011).

### Participants

In accordance with the aims of this study, a subset of patients who were likely to have more severe cognitive impairment was chosen and comprised of those diagnosed with dementia. This study evaluated a group of healthy controls (*n*=132) and patients diagnosed with dementia (*n*=191). Each group was randomised and further split into a ‘training’ cohort (70%) for development of the short-form NUCOG and a ‘testing’ cohort (30%) for validation of the shortened versions.

### Item selection

All statistical analysis was performed using IBM SPSS Version 29 and Medcalc Software Version 23.1.7. Receiver operating characteristic (ROC) curves were computed from data belonging to the ‘training’ cohort to determine the NUCOG items that best discriminate between healthy controls and patients with dementia. The area under the curve (AUC) was calculated for each item of the original NUCOG. The higher the AUC, the better the ability of the item to discriminate between groups, with an AUC above 0.9 deemed excellent, 0.8-0.9 good, 0.7-0.8 poor, 0.6-0.7 worthless and 0.5-0.6 failed (Metz, 1978). The items were ranked by AUC, generating a top-5, top-10 and top-15 list. Additional items were added to each short-form NUCOG for functionality reasons i.e. C1reg (registration) was included to make C1rec (verbal recall) work in the 15-item version and 10-item versions; B1 (figure copy) was included to make C2 (spatial recall) work in the 5-item and 10-item versions. The selected items for each short-form version can be seen in Table 1. Although each short-form NUCOG contains one or two additional items, they will continue to be referred to as 5-item, 10-item and 15-item for ease, reflecting the core items identified in each analysis.

**Table 1.**
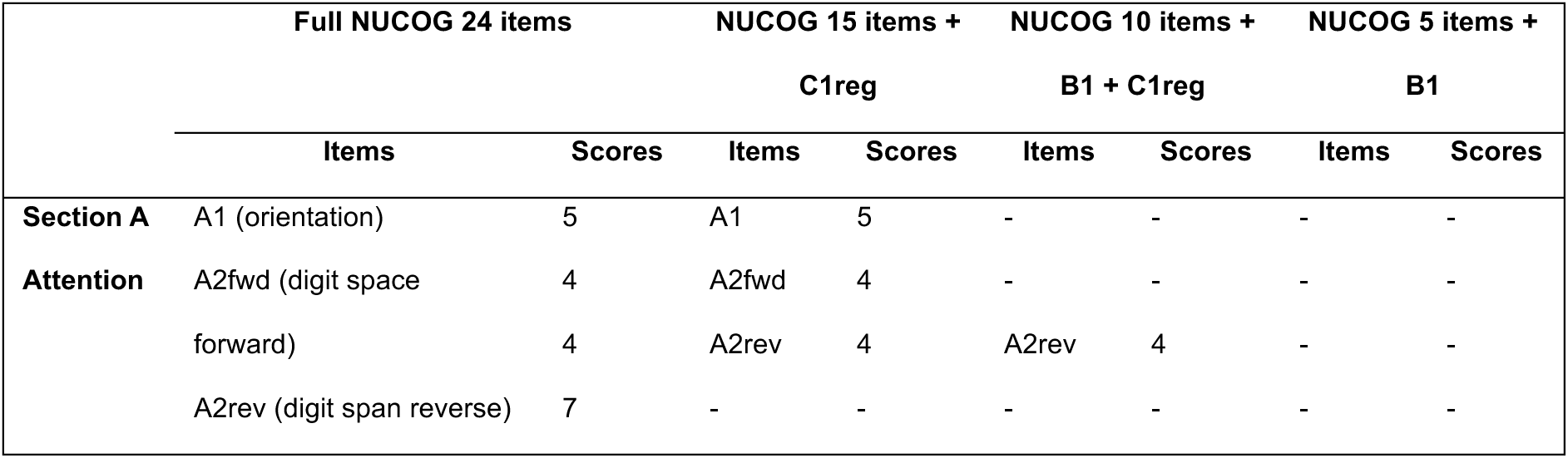
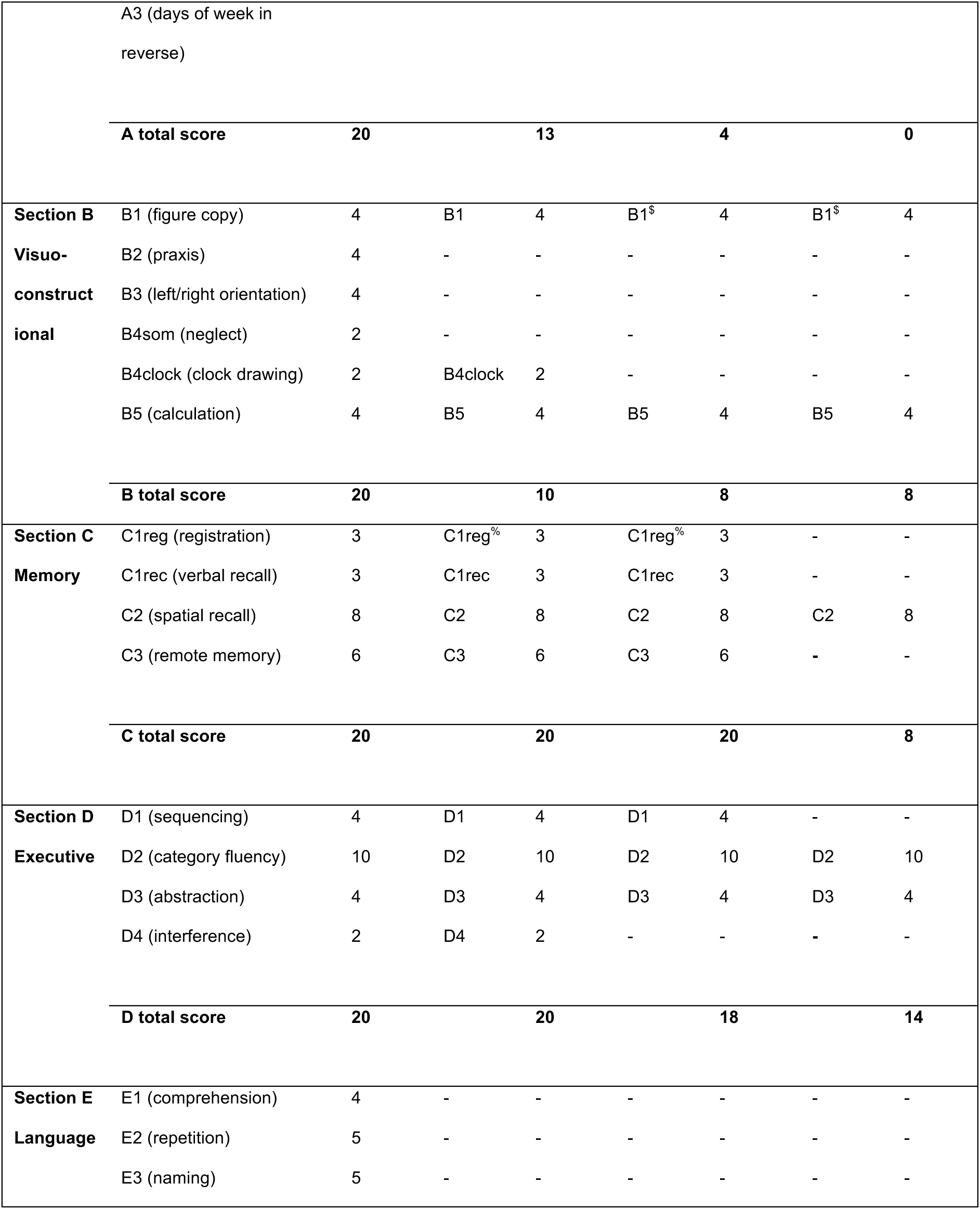
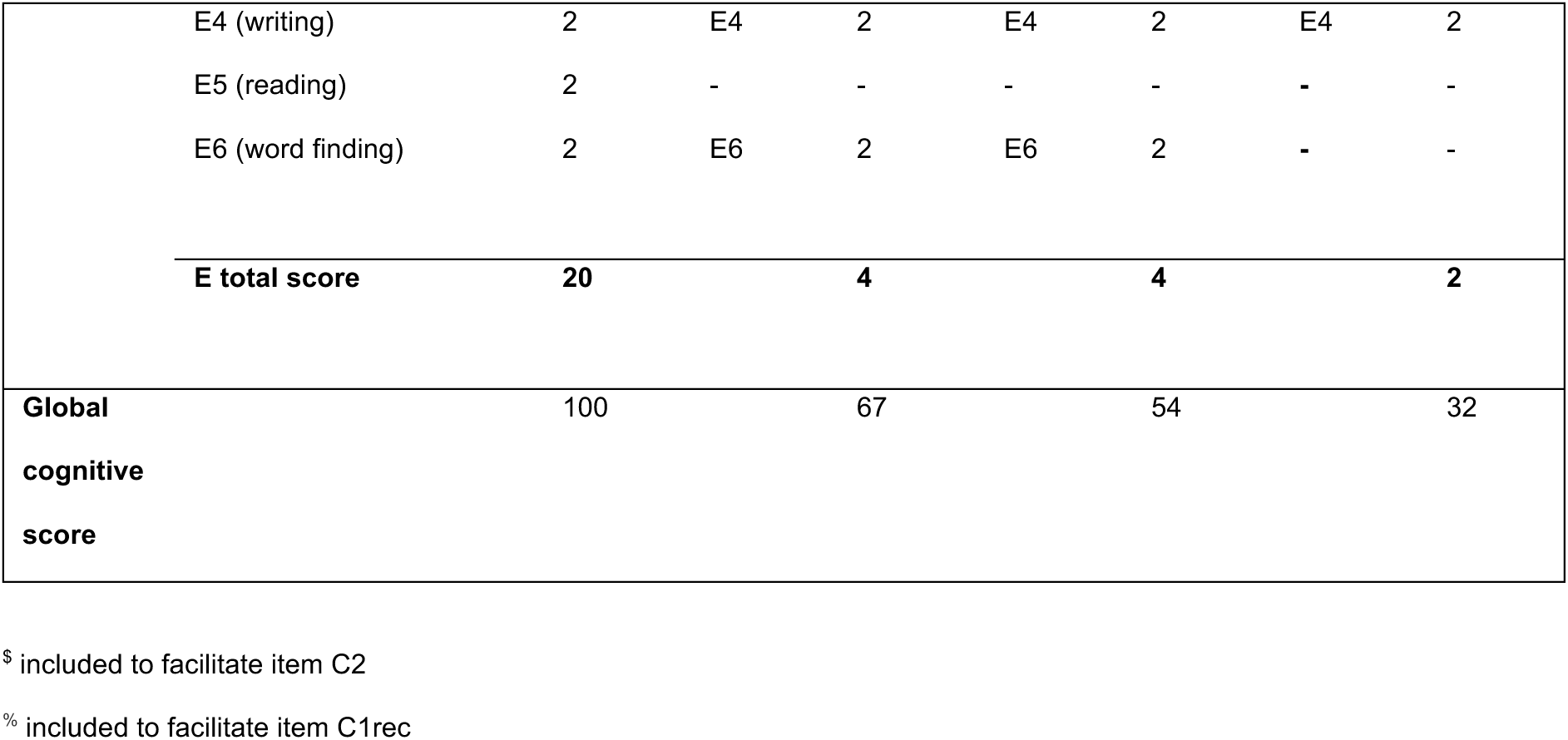
Versions of short-form NUCOG in comparison with original NUCOG

### Statistical analysis

The psychometric properties of each short-form version were examined independently to the original NUCOG. Criterion validity was tested by examining the AUC of the global cognitive function score for each of the NUCOG versions, including the original NUCOG as a gold standard comparison. The optimal cut-off value was derived from the ‘training’ cohort using Youden’s index and applied to the ‘testing’ cohort to generate sensitivities, specificities, positive predictive values (PPV), and negative predictive values (NPV) for validation.

Independent samples t-tests were performed on the global scores to examine mean differences between diagnostic groups. Standardised effect sizes were computed as Cohen’s *d*. Convergent validity was calculated via Kendall’s tau-b correlation coefficient between the original and new NUCOG global scores. Psychometric reliability was examined using Cronbach’s α.

## Results

### Clinicodemographic characteristics

This study included 323 individuals, of which 132 were healthy controls (41%) and 191 were diagnosed with dementia (59%). Demographic characteristics are summarised in Table 2. Among those with dementia, the mean age was 61.05 (*SD*=10.78), with ages ranging from 24 to 88 years. Most patients with dementia were aged 50 or above (*n*=172, 90%). There were more females than males in the control group (53%), while males made up the majority in the dementia group (60%). Most of the retrospective data on patients with dementia were sourced from the Neuropsychiatry inpatient unit at the Royal Melbourne Hospital (*n*=138, 72%), while controls were from Melbourne (*n*=89, 67%) and Brisbane (*n*=29, 22%), as described in the Methods section. The controls completed more years in education and scored higher in all NUCOG domains – the mean global score for controls was 92.78 (*SD*=4.89) in comparison to 61.03 (*SD*=17.71) in the dementia group.

**Table 2.** Comparison of characteristics between training and testing groups in both control and dementia groups (*N* = 323)

| *Characteristics* | Controls group |  |  | Disease group (Dementia) |  |  |
| --- | --- | --- | --- | --- | --- | --- |
| | Training Group<br>$n = 92$ (70%) | Testing Group<br>$n = 40$ (30%) | Total<br>$n = 132$ | Training Group<br>$n = 134$ (70%) | Testing Group<br>$n = 57$ (30%) | Total $n = 191$ |
| *Age*<br>[mean (SD) years] | 49.93 (19.05)<br>Range: 18 - 93 | 49.00 (17.97)<br>Range: 20 - 75 | **49.65**<br>**(18.67)** | 60.38 (11.10)<br>Range: 24 - 82 | 62.63 (9.91)<br>Range: 33 - 88 | **61.05 (10.78)** |
| *Gender* |  |  |  |  |  |  |
| Male | 44 (48%) | 18 (45%) | **62 (47%)** | 78 (58%) | 36 (63%) | **114 (60%)** |
| Female | 48 (52%) | 22 (55%) | **70 (53%)** | 55 (41%) | 21 (37%) | **76 (40%)** |
| Other | - | - | - | 1 (<1%) | - | **1 (<1%)** |
| *Education* | 14.04 (3.12) | 13.88 (2.51) | **13.99 (2.94)** | 11.36 (3.62) | 11.51 (3.45) | **11.40 (3.55)** |
| *[mean (SD) years]* |  |  |  | (Missing data<br>= 15; n = 119) | (Missing data<br>= 10; n = 47) | **(Missing**<br>**data = 25, n**<br>**= 166)** |
| *Setting* |  |  |  |  |  |  |
| NPU Inpatient | 3 (3%) | - | **3 (2%)** | 97 (72%) | 41 (72%) | **138 (72%)** |
| NPU | 4 (4%) | 2 (5%) | **6 (5%)** | 18 (13%) | 11 (20%) | **29 (15%)** |
| Outpatient |  |  |  |  |  |  |
| NPU CL | - | - | - | 2 (1%) | 2 (4%) | **4 (2%)** |
| Private | 1 (1%) | 1 (3%) | **2 (2%)** | 12 (9%) | 2 (4%) | **14 (7%)** |
| Practice |  |  |  |  |  |  |
| Acute IPU | - | - | - | 4 (3%) | - | **4 (2%)** |
| Original | 60 (65%) | 29 (73%) | **89 (67%)** | - | - | - |
| Controls |  |  |  |  |  |  |
| PAH Controls | 21 (23%) | 8 (20%) | **29 (22%)** | - | - | - |
| Missing data | 3 (3%) | - | **3 (2%)** | 1 (1%) | 1 (2%) | **2 (1%)** |
| *NUCOG Global* | 92.60 (5.07) | 93.20 (4.47) | **92.78 (4.89)** | 61.98 (17.42) | 58.79 (18.34) | **61.03 (17.71)** |
| *Score [mean (SD)]* |  |  |  |  |  |  |
| *A total score* | 17.64 (1.57) | 17.18 (2.09) | **17.50 (1.75)** | 12.37 (4.68) | 11.56 (4.74) | **12.13 (4.70)** |
| *B total score* | 19.08 (1.41) | 19.55 (0.78) | **19.22 (1.27)** | 14.36 (4.03) | 13.96 (4.31) | **14.24 (4.11)** |
| *C total score* | 18.07 (2.20) | 18.55 (1.52) | **18.21 (2.03)** | 10.36 (4.47) | 9.12 (4.10) | **9.99 (4.39)** |
| *D total score* | 17.91 (2.54) | 18.00 (2.17) | **17.94 (2.43)** | 9.34 (4.39) | 8.90 (4.71) | **9.21 (4.48)** |
| *E total score* | 19.76 (0.51) | 19.70 (0.62) | **19.74 (0.54)** | 15.71 (3.43) | 15.24 (3.92) | **15.57 (3.58)** |

**Abbreviations:** NPU = Neuropsychiatry Unit, CL = consultant-liaison, IPU = inpatient unit, PAH = Princess Alexandra Hospital

Table 3 illustrates the distribution of dementia subtypes within the dementia group as well as across the training and testing cohorts. Of the 191 dementia patients, the most common dementia diagnosis was Alzheimer’s disease (*n*=54, 28%), followed by frontotemporal dementia (*n*=53, 28%). Other aetiologies included dementia not otherwise specified (NOS) (*n*=22), vascular dementia (*n*=18), Korsakoff’s/alcoholic dementia (*n*=10), dementia with lewy bodies (DLB) (*n*=8), posterior cortical atrophy (*n*=7), Parkinson’s disease dementia (*n*=6), semantic dementia (*n*=4), progressive supranuclear palsy (PSP) (*n*=4) and dementia secondary to other causes (*n*=5).

**Table 3.** Dementia subgroups among training and testing groups (*n*=191)

| Dementia Diagnosis | Disease Group (Dementia) |  |  |
| --- | --- | --- | --- |
| | Training Group<br>$n=134$ | Testing Group<br>$n=57$ | Total<br>$n=191$ |
| Alzheimer's disease | 39 (29%) | 15 (27%) | **54 (28%)** |
| Frontotemporal dementia (FTD) | 43 (33%) | 10 (18%) | **53 (28%)** |
| Dementia Not Otherwise Specified (NOS) | 11 (8%) | 11 (19%) | **22 (12%)** |
| Vascular dementia | 13 (10%) | 5 (9%) | **18 (9%)** |
| Korsakoff's/Alcoholic dementia | 6 (4%) | 4 (7%) | **10 (5%)** |
| Dementia with Lewy Bodies (DLB) | 6 (4%) | 2 (4%) | **8 (4%)** |
| Posterior Cortical Atrophy | 6 (4%) | 1 (2%) | **7 (4%)** |
| Parkinson's disease<br>dementia | 3 (2%) | 3 (5%) | **6 (3%)** |
| Semantic dementia | 1 (1%) | 3 (5%) | **4 (2%)** |
| Progressive<br>supranuclear palsy<br>(PSP) | 1 (1%) | 3 (5%) | **4 (2%)** |
| Dementia secondary to<br>other causes | 5 (4%) | - | **5 (3%)** |

### Short-form NUCOG versions

ROC curves were created for each of the 24 items and were ranked by their AUC value, as demonstrated in Figure 1. Eight items (D2, C2, E4, D3, B5, A2rev, C3, E6) had an AUC value above 0.8. Based on the calculated AUC values, three separate short-form NUCOG tests were created – 15-item + C1reg; 10-item + B1 + C1reg; and 5-item + B1 versions (Table 1). The AUC of the global scores for each version were then calculated.

**Figure 1.**
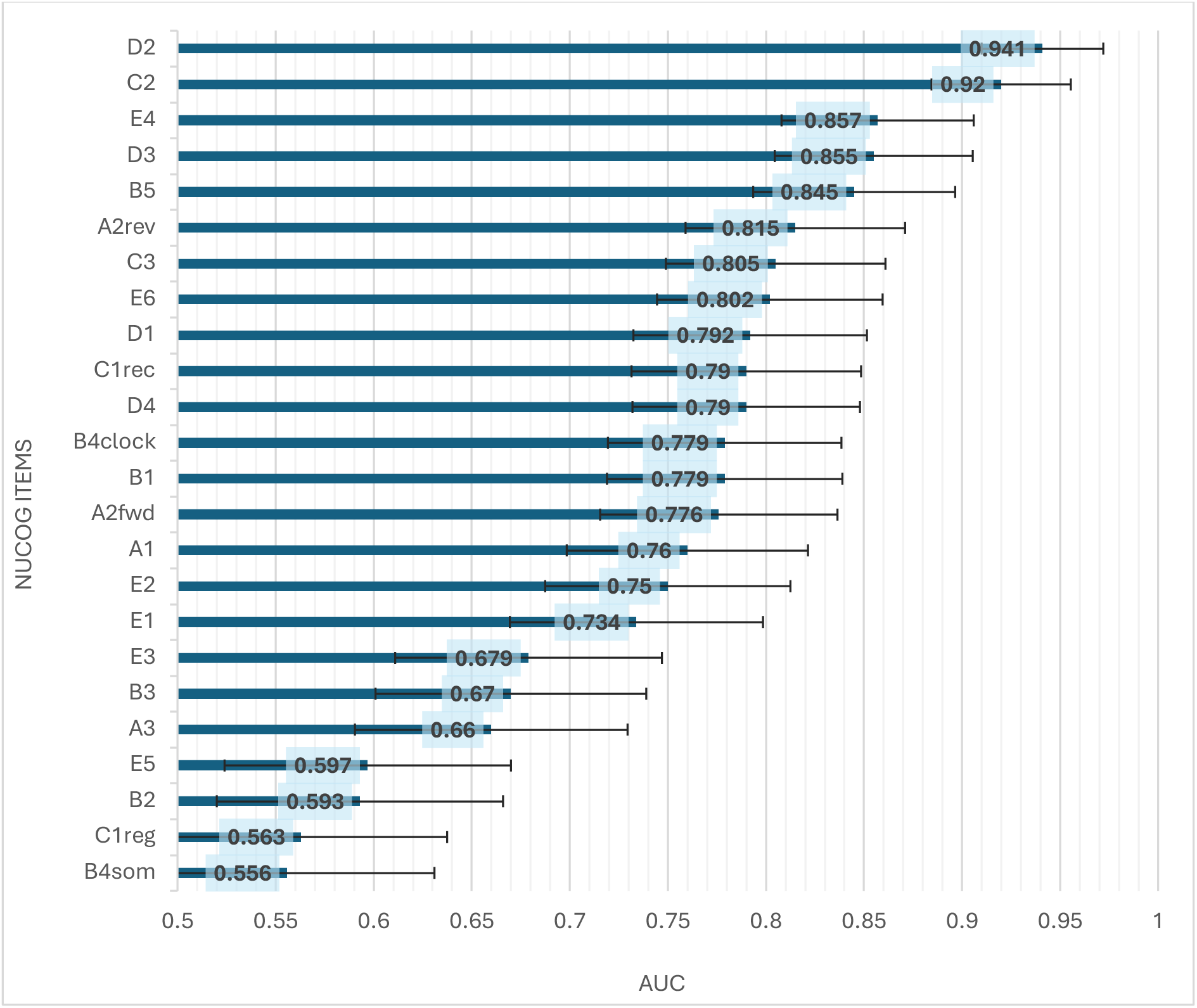
24-items of NUCOG in rank order based on AUC

The AUC of the total scores were then calculated initially using the ‘training’ cohort. The optimal cut-off scores were generated using Youden’s index as seen in Table 4. Subsequently, validation was performed using the ‘testing’ cohort. Using the optimal cut-off score of 85/100 in this study, the sensitivity and specificity of the original NUCOG for the detection of dementia was 0.95 and 0.98 respectively, with a PPV of 0.98 and NPV of 0.93. In comparison, the 10-item version, with a cut-off score of 42/54, generated similar values as the original NUCOG, with a sensitivity of 0.98, specificity of 0.95, PPV of 0.97 and NPV of 0.97. The sensitivity reduced as the tests contained less items, however the 5-item NUCOG still maintained a sensitivity of 0.91 and specificity of 0.98 at a cut-off score of 24.5/32.

**Table 4.** Performance of each NUCOG version in comparison to the gold-standard full NUCOG

|  |  |  | Training cohort (70%), *n*=226 |  |  |  |  | Testing cohort (30%), *n*=97 |  |  |  |  |
| --- | --- | --- | --- | --- | --- | --- | --- | --- | --- | --- | --- | --- |
| Test | Youden's Index | Cut-off score | AUC [95% CI] | Sensitivity [95% CI] | Specificity [95% CI] | PPV [95% CI] | NPV [95% CI] | AUC [95% CI] | Sensitivity [95% CI] | Specificity [95% CI] | PPV [95% CI] | NPV [95% CI] |
| Full NUCOG | 0.868 | 85/100 | 0.981<br>[0.967, 0.996] | 0.96<br>[0.91 - 0.98] | 0.91<br>[0.84 – 0.96] | 0.94<br>[0.89 - 0.97] | 0.93<br>[0.86 – 0.97] | 0.996<br>[0.988, 1.003] | 0.95<br>[0.85 – 0.99] | 0.98<br>[0.87 – 1.00] | 0.98<br>[0.88 – 1.0] | 0.93<br>[0.81 – 0.98] |
| 15 items + C1reg | 0.876 | 54/67 | 0.980<br>[0.965, 0.996] | 0.96<br>[0.92 - 0.99] | 0.91<br>[0.84 – 0.96] | 0.94<br>[0.89 – 0.97] | 0.94<br>[0.88 – 0.98] | 0.996<br>[0.990, 1.003] | 1.00<br>[0.94 – 1.00] | 0.70<br>[0.53 – 0.83] | 0.83<br>[0.75 – 0.88] | 1.00<br>[0.88 – 1.00] |
| 10 items + B1 + C1reg | 0.872 | 42/54 | 0.981<br>[0.966, 0.996] | 0.95<br>[0.90 – 0.98] | 0.92<br>[0.85 – 0.97] | 0.95<br>[0.90 – 0.97] | 0.92<br>[0.85 – 0.96] | 0.997<br>[0.991, 1.003] | 0.98<br>[0.91 – 1.00] | 0.95<br>[0.83 – 0.99] | 0.97<br>[0.88 – 0.99] | 0.97<br>[0.84 – 1.0] |
| 5 items + B1 | 0.856 | 24.5/32 | 0.979<br>[0.965, 0.993] | 0.91<br>[0.85 – 0.95] | 0.95<br>[0.88 – 0.98] | 0.96<br>[0.91 – 0.98] | 0.88<br>[0.80 – 0.98] | 0.993<br>[0.982, 1.003] | 0.91<br>[0.81 – 0.97] | 0.98<br>[0.87 – 1.00] | 0.98<br>[0.88 – 1.00] | 0.89<br>[0.77 – 0.95] |

### Group differences

In the ‘training’ cohort, the mean global NUCOG score for controls (*n*=92, *M*=92.60, *SD*=5.07) was significantly higher than dementia patients (*n*=134, *M*=61.98, *SD*=17.43, *t*(224)=16.37, p < 0.001. The mean difference was 30.62, 95% CI [26.93, 34.30]. The effect size statistic, *d*=2.22, 95% CI [1.89, 2.55] indicates that the mean global NUCOG score for controls is 2.22 standard deviations higher than dementia patients. The short-form versions all have higher effect sizes than the original NUCOG, with Cohen’s *d* ranging from 2.48, 95% CI [2.13, 2.83] to 2.59, 95% CI [2.23, 2.95]. This trend was also reflected in the ‘testing’ cohort, as illustrated in Table 5. Furthermore, each mean difference between the two variables were statistically significant (p < 0.001).

**Table 5.** Effect size measuring mean difference in global NUCOG scores between control and dementia groups

|  |  | Training Cohort (70%) |  | Testing Cohort (30%) |  |
| --- | --- | --- | --- | --- | --- |
| Test | Global score | Mean Difference [95% CI] | Cohen's *d* [95% CI] | Mean Difference [95% CI] | Cohen's *d* [95% CI] |
| Full NUCOG | 100 | 30.62 [26.94, 34.30] | 2.22 [1.88, 2.55] | 34.41 [28.53, 40.29] | 2.40 [1.86, 2.92] |
| 15 items + C1reg | 67 | 25.55 [22.80, 28.30] | 2.48 [2.13, 2.83] | 27.44 [23.70, 31.17] | 3.01 [2.42, 3.59] |
| 10 items + B1 +<br>C1reg | 54 | 21.36 [19.14, 23.58] | 2.57 [2.21, 2.92] | 24.13 [20.91, 27.35] | 3.07 [2.47, 3.66] |
| 5 items + B1 | 32 | 14.46 [12.97, 15.95] | 2.59 [2.23, 2.94] | 15.37 [13.19, 17.54] | 2.89 [2.31, 3.47] |

### Convergent validity

Kendall’s tau-b correlation coefficients reflected very high correlation between the global scores of the original NUCOG and short-form versions: 0.95 with the 15-item version, 0.92 with the 10-item version and 0.85 with the 5-item version.

### Reliability

Psychometric reliability of the original NUCOG measured by Cronbach’s *α* was 0.93. The Cronbach’s *α* remained above 0.80 for the shortened versions of NUCOG: 0.92 in 15-item version; 0.90 in 10-item version; 0.83 in 5-item version.

## Discussion

All three abbreviated versions of the NUCOG showed high reliability and validity, and have similar psychometric properties to the original, with the added benefit of shortened administration time; the 15-item version takes around 15 minutes, 10-item version takes 10 minutes and the 5-item version takes 5 minutes. In the original validation paper of the NUCOG, the sensitivity for the detection of dementia was 0.84, and the specificity was 0.98 at a cut-off score of 80/100 (Walterfang et al., 2006). In this study, a higher cut-off score of 85/100 yielded higher sensitivity at 0.95 and same specificity 0.98. Incorporating the top items that have the highest discriminatory validity reduces the likelihood of ceiling effects, as demonstrated by the robust sensitivity values such as 0.98 in the 10-item version (refer Table 1 for items in each version).

Two items had an AUC above 0.9, embodying excellent discriminatory function in these items between normal controls and those with dementia: categorical fluency (D2), and spatial recall (C2). The other items included in each short-form version include writing a sentence (E4); abstraction (D3); and calculation (B5). This is contrary to findings that the best discriminatory items between dementia and controls in both MMSE and MOCA include orientation, word recall and serial subtraction, which have been included in their respective short-form cognitive tests (Haubois et al., 2011; Horton et al., 2015; Schultz-Larsen et al., 2007).

Very brief cognitive screening tests have their greatest utility in primary care settings, where time is most limited – among those identified as efficient, well-accepted by clinicians, and minimally affected by sex, education and race, include the General Practitioner Assessment of Cognition (GPCOG), Mini-Cog and Memory Impairment Screen (MIS) (Brodaty et al., 2006; Milne et al., 2008). The Mini-Cog is a very brief screening tool which takes three minutes to administer and contains three items: three-word registration, clock drawing and three-word recall, conceptualised as a measure of ‘cognitive vital signs’ (Borson et al., 2000). According to a meta-analysis of six studies which used Mini-Cog as a screening test in patients ≥ 60 years old, it was found to have a comparatively lower sensitivity of 0.76 and specificity of 0.83 in detecting dementia (Abayomi et al., 2024). Alternatively, the MIS is a four-item cognitive test which takes four minutes to administer, and focuses on memory with delayed free and cued recall (Buschke et al., 1999). According to a systematic review by Yokomizo et al. (2014), its sensitivity ranges from 0.74 – 0.86 and specificity ranges from 0.96 – 0.97, however a disadvantage with this test is that there is no direct measure of executive function or visuospatial ability, limiting its applicability in detecting non-amnesic mild cognitive impairment and other types of dementia beyond Alzheimer’s disease. Despite only containing a very limited number of items, the 5-item NUCOG version contains an item from each of the five original cognitive domains, except for attention, demonstrating its breadth within its rapid assessment in comparison to other cognitive tests of similar length.

A comparable short-form cognitive test that takes five minutes to administer and covers a broader range of cognitive domains is the Mini-Addenbrooke’s Cognitive Examination (M-ACE), which tests orientation, memory, verbal fluency and visuospatial function using five items (Hsieh et al., 2014). This test has been shown to be more sensitive, with somewhat less specificity, at all cut-off points compared to the MMSE (Hsieh et al., 2014). The only overlapping category is its language fluency test with the D2 test (categorical fluency) on the NUCOG; the 5-item NUCOG may provide a limited cognitive profile and perhaps a longer screening test may increase the ability to differentiate between different pathologies.

An advantage of 10-item version is that it contains items that test executive functioning broadly through sequencing (D1), category fluency (D2) and abstraction (D3), and both declarative semantic memory (C3) and episodic memory through both verbal recall (C1) and spatial recall (C2). Executive functioning and spatial recall are neglected in other cognitive tests such as the MMSE, which may yield scores in the ‘normal’ range, even in the presence of severe functional limitations or executive dysfunction (Juby et al., 2002). Furthermore, the NUCOG contains a spatial recall item (C2) which has been shown to be able to differentiate between Alzheimer’s disease from non-dementia disease, while constructional praxis copy, which is in the MOCA, cannot (Li et al., 2023). Although the 15-item version retains orientation and clock drawing test, the 10-item NUCOG (“NUCOG10”) appears to provide an ideal balance of duration to administer with very high sensitivity and specificity, very high validity, and the retention of items from each cognitive domain from the long-form test.

## Limitations

Data was largely derived from patients from a specialised tertiary service, often with cognitive disorders, limiting generalisability to the general population. For the purposes of statistical analysis, the diagnostic classification was oversimplified, with patients assigned a single primary diagnosis. As befits patients from a tertiary referral service, most had other comorbid conditions, including neurological or psychiatric disorders. Even if these conditions contributed cognitive impairment (such as, for example, major depression), if patients had a primary dementia, this was their primary diagnosis.

Even though some patients were tested on multiple occasions, only one NUCOG was recorded - either the first one performed or the one closest to when the primary diagnosis was made. As such, the time between the NUCOG and definitive diagnosis varied. Not only were clinicians not blinded to the NUCOG score when making the diagnosis, but the NUCOG contributed to the diagnostic process, which may account for the significant differences between the scores between the control and dementia groups. As the data spans 23 years from 2001 to 2023, the diagnostic criteria or diagnostic process for dementia may have varied over time.

Another limitation of this study is that the dementia group was treated as a homogenous group in the statistical analysis and the ability to differentiate between dementia subtypes or those with mild cognitive impairment was not explored. No adjustments were made for age, education, and sex, which may have confounded results.

## Conclusion

This study introduces three abbreviated versions of NUCOG, which provide clinicians with a time-efficient cognitive screening tool without compromising reliability or validity. The 10-item version of the NUCOG appears to offer an ideal balance between breadth and brevity of cognitive testing, maintaining classification accuracy with sensitivity, specificity and predictive values for dementia on par with those of the original NUCOG. Further directions would involve testing these short-form versions for performance in non-tertiary cohorts, across dementia subtypes and in non-dementia groups.

## Data Availability

The datasets generated during and/or analysed during the current study are available from the corresponding author on reasonable request.

## Acknowledgements

The authors would like to gratefully acknowledge the contribution to the control group by A/Prof Frances Dark from her unpublished dataset from Brisbane, Australia.

## Author contributions

MW, SL, CM and AL contributed to the design of the research. All authors were involved in the data collection. Statistical analysis was conducted by AL and CM. AL wrote the first draft of the manuscript, and all authors contributed to and approved the final manuscript for submission.

## Statements and declarations

### Ethical considerations

Prospective and retrospective ethics have been obtained for data used in this research, as approved by the Royal Melbourne Hospital Human Research Ethics Committee (2005.198; 2018.371; and 2020.142).

